# Aging in Place - A mixed methods study protocol of how care providers in Sweden organise and adapt granted home care services to the preferences and needs of people living with dementia

**DOI:** 10.64898/2026.04.29.26352070

**Authors:** Annika Taghizadeh Larsson, Anna Olaison, Lars-Christer Hydén, Mattias Antar, Ceciel Heijkants, Jimmy Lindberg, Sofia Nordmark, Veronika Wallroth, Susanne Kelfve

**Affiliations:** Department of Culture and Society, Division of Social Work, Linköping University, Sweden; Department of Culture and Society, Division of Ageing and Social Change, Linköping University, Sweden

## Abstract

The project *Aging in Place* applies a novel approach to investigate how social care services in Sweden are adapted to preferences and needs of older persons living at home with dementia (including their partners). The project covers the process that starts when a person is granted to receive services, which are communicated to a care provider – who should organize the services – and finally reach the receiving older person. This approach differs from previous research on elder care, which has focused either on the purchaser or the provider side of Swedish municipal elder care in the purchaser-provider model Sweden adopted since the mid-1990s. The project focuses on: 1) what specific social care services older persons living at home with dementia are granted; if a dementia diagnosis is needed for some services; and the differences between municipalities; 2) how care providers organize granted services and adapt them to people living with dementia; including differences between municipalities and care provider units; 3) how care recipients living with dementia (including co-habiting partners) experience and influence the receiving of services. The ambition is to generate both generalizable knowledge about social care services for people living with dementia in Sweden, including differences between municipalities and care providers, and in-depth, exploratory knowledge about how care providers organise services and how these services are received by care recipients. The project encompasses an innovative, and necessary, combination of methods and materials: register studies, web surveys, as well as observations and interviews. The project will provide important, elemental, knowledge on Swedish dementia- and eldercare. This knowledge is needed as a basis for further studies and as a contribution to discussions on how future social care can be developed to ensure people living with dementia, and their partners, equality, participation, and dignity in later life.

## Introduction

Dementia is a major public health problem that affects individuals and families all over the world and causes increasing challenges at the societal level. The growing number of people living with dementia, particularly Alzheimer’s disease, poses challenges to the organization of care, the supply of staff, and staff education. Therefore, there is an urgent need to expand our understanding of the organization and quality of dementia care, how individuals with dementia experience receiving support, and the type of support they request.

Recent studies indicate that the percentage of people in Sweden diagnosed with dementia who receive home care is as large as the percentage who live in residential care, or more specifically, in special housing (1). Despite this, knowledge about social care aimed at people with dementia in their own homes is limited. This project meets this research gap by generating generalizable knowledge of social care services for people living with dementia in Sweden, including differences between municipalities and home care providers, and in-depth, exploratory knowledge of the organisation of services by providers and finally how services are received by care recipients.

The project covers the process from granted to received social care services, including how (and if) services are adapted, and (re)designed to meet the particular needs and preferences of older people with cognitive and communicative impairments due to dementia, in several steps as part of this process. In addition, the project includes the voices of the older persons with dementia and their co-habiting partners on how current services are perceived and how they prefer the services to be provided.

The project has a societal, scientific, and international relevance because it addresses one of the most important future areas of social welfare: how can aging in place be achieved to ensure people living with dementia and their partners, equality, participation, and dignity in later life?

### Provision of social services in the Swedish eldercare system

In formal terms and at the national level, the social care services that older adults generally receive in Sweden are regulated under the Social Service Act (SSA) and aims to ensure a reasonable standard of living. Care services are tax-subsidised and provided to eligible citizens through a needs assessment process (2). The municipality is responsible for the provision of social care and is the formal decision-making authority when it comes to granting services. It is also a municipal task, and freedom, to decide what a reasonable standard of living specifically implies on the local level for its citizens with care needs, given the specific conditions for each municipality. In the mid-1990s, a purchaser provider system, including a system of care management, was introduced for care services, with the stated intention of facilitating the coordination of support for older people and increase legal certainty (3) (4). Then, in 2009, the Act on System of Choice in the Public Sector was introduced (in Swedish: *Lagen om valfrihetssystem* [LOV]) which made it possible – but not mandatory – for municipalities to expose the execution of granted services to market competition, and to enable users to choose among providers. Consequently, in today’s Sweden, the investigation of individual needs for social care services to people with dementia in ordinary housing is carried out by a municipal needs assessor (in Swedish: *biståndshandläggare*), often a social worker, and a decision on granted care, an “order” (in Swedish: *beställning*), is handed over to a care provider; formally chosen by the older person among several available providers, or given without choice, depending on if the municipality in question has implemented LOV, or not.

### State of the art

The vast social scientific research on home care carried out in Sweden since the 1990s has focused on care for older people in general (5) (6). During the last decade, there has been a growing scholarly interest in the role of needs assessors, that is, the persons who investigate older people’s need of social care services before the decisions on granted services are handed over to social care providers in the purchaser-provider system of the Swedish elder care (7) (8) (9). What specific care services older people living with dementia are granted, and what information the needs assessors transfer to the social care providers after a decision on social care is made, is largely unknown. Neither do we have knowledge about how social service providers organise the specific granted services for older people (both for those with and without dementia) living in ordinary housing, or how the services are experienced by the older persons themselves and their relatives. There is also limited knowledge about how the organisation, type, and amount of care might differ within and between municipalities. Some municipalities have for instance a special dementia team for the provision of care, and many municipalities use several different care service providers (both private and municipal), especially in the big cities (1) (10).

Internationally, studies show that older persons with dementia are less likely to be invited to engage in their home care compared to those without dementia (11) (12). Older persons with dementia receiving home care services highlight the importance of values of reciprocity, respect, attentiveness, and safety in care situations (13). Also, being treated as a unique and competent human, despite the dementia diagnosis and in need of support, are important factors. Studies also indicated that persons living with dementia and their relatives are frustrated with the inconsistent staff rostering in home care and are requesting more specific dementia knowledge from care workers (14). Consistent with these results, studies carried out in Sweden have shown that the quality of home care services is dependent on the personal relationship between the care worker and the care recipient. High staff continuity, flexibility, i.e., space for the care worker to adapt the provision of care to the individual’s personal and varying situation and needs, and sufficient time for staff to handle individual needs and unexpected situations, have been highlighted as necessary factors for home care to meet the requirements of the Social Services Act’s value base. Further, good competence and opportunities for care workers to obtain further training, supervision and reflection have been described as important. When it comes to increasing the prerequisites for people with dementia to continue to live safely in their homes, specialized knowledge among care workers have been highlighted as important (15).

Family members are often the main care provider for persons living with dementia at home (16) (17). Dementia can have consequences for couples where there is an increased risk for burden, stress, depression, and other health complications for the cohabiting partner (18). Still, having a partner with dementia does not always mean only negative things. The strong sense of togetherness between partners also helps them to face life’s hardships (19). This ongoing feeling of togetherness, despite change, makes the family situation somewhat ambivalent because of the many practical, emotional, and moral pressures involved (20).

Research showed that home care workers specialized in dementia care report higher job strain than other home care workers (21). Staff in dementia care also tend to focus more on physical safety and minimizing risk (22) (23). They also face ethical dilemmas in balancing between reducing possible risks and protect persons with dementia and at the same time consider the individual’s autonomy (24). A few studies have investigated how risks for persons with dementia living at home can be handled in actual care situations where there is a co-habiting relative, and how risks might influence how care is performed in the everyday life of older persons with dementia living at home (21).

In sum, previous international and Swedish studies have investigated social services for persons with dementia from different perspectives, either from the older persons, relatives, or the home care worker. However, none of these studies have focused specifically on the care process (18) (25) (encompassing how the services provided within elder care are adapted to needs related to dementia as part of designing and carrying out the services – or how the recipients, including co-habiting partners, experience and can influence the services and how they want services to be designed. Although there is some national statistics about social care services granted and organised in Sweden, little is known about elemental issues in focus of this project, such as:

- Is a dementia diagnosis required to access certain types of social care services?
- What information regarding the needs of individuals living with dementia (and cohabiting partners) is transferred from the needs assessor to the care provider?
- Who is responsible for designing and planning the granted care services within the care-providing organisation?
- Are the granted services adapted to the older person’s cognitive abilities?
- Are there any differences in the care services provided to people with dementia across municipalities or between different care providers?
- How are the care services delivered during actual home visits by home-care staff?
- How do older persons with dementia and their relatives experience, and exert influence over, the care provided?

## Materials and Methods

In this innovative project we will use a mix of register data, questionnaires, interviews, and observations to capture the process from a granted decision to the actual receiving of social care services, including the experience of the older person with dementia and their relatives.

### The project has three specific aims

(a) to map and describe the provision of social care services (in Swedish *insatser*) to people living with dementia in ordinary housing in Sweden; (b) to explore how these social care services are adapted by the service providers to the needs and preferences of older persons with dementia; and how the services are experienced by the person with dementia themselves and their cohabiting partners; and (c) to compare the provision and the adaptation of the granted services between municipalities and social care providers.

#### The project consists of three sub studies

##### Sub-study 1 – An overall and comparative picture of social care services available for and granted to people with dementia living in ordinary housing

The overall aim of this sub-study is to map the variety of social care services to people living with dementia in ordinary housing in Sweden’s 290 municipalities, by describing a) what social care services people with dementia are granted (register data), b) what specialised social care services adapted to people with dementia are available in municipalities (survey data), c) if a dementia diagnosis is a criterion for some of the services (survey data), and d) if there are any differences between municipalities regarding a, b and c?

Two different data sources will be used. First, we will collect survey data by sending out a web questionnaire to needs assessors in all municipalities in Sweden with questions about what specialized services designed to meet the needs of people with dementia the municipalities offers (e.g. dementia teams or specific adapted services such as so called *boendeservice*) and if some of these services require a dementia diagnosis. Second, we will use register data stored at the Division of Ageing and Social Change, Linköping University, including the National in- and outpatient registers and the Social Service Register (latest updated 2024). The national in- and outpatient registers will be used to identify people with a dementia diagnosis and with the Social Service Register we will identify what social care services people with a dementia diagnosis are granted. All analyses will be stratified by municipality.

The second data source, the web survey, will give information about available specialised social care services in the municipalities as well as differences between municipalities in this respect. With the combination of the register data (information about what social care services people with a dementia diagnosis are granted) and the web survey (insight in available specialised services for people with dementia), we are able to evaluate to what extent the reported available specialised services are reflected in the granted services to people with a dementia diagnosis.

All data will be described and analysed using STATA 17 (StataCorp, College Station, TX) using common statistical methods, such as frequencies, proportions, Chi2, t-test, and Anova, depending on outcome variable and research question. For the description of available specialised social care services in the municipalities, simple proportions will primarily be used, while comparisons between municipalities will primarily be analysed by e.g., Chi2, t-test, and Anova.

##### Sub-study 2 -The organisation of granted social care services by home care providers

Sub-study 2 aims to explore, describe and compare the organisation of granted social care services by home care providers to people living with dementia in ordinary housing. Focus is on how home care providers in Swedish municipalities plan and carry out granted services and whether and how providers (managers and care workers) consider and adapt to needs related to dementia, and individual preferences of care recipients living with dementia (including cohabiting partners), when the services are organised.

The sub-study consists of digital (zoom) interviews with managers and care workers and will entail both closed and open-ended questions. The closed-ended questions address the organisation and adaption of social services. This includes the level of staff continuity (in Swedish *personalkontinuitet*); staff education, frequency and duration (i.e. how often services are carried out and for how long care workers stay in the home of/with the person each time), and flexibility (i.e. to what extent the content of a home visit and the time the care worker will spend together with the person with dementia is flexible when the care worker enters the home of a care recipient). Further, the overall type of information included in the order from the purchaser-side in the municipalities of Sweden will be mapped and compared, as will transferred information regarding dementia diagnosis, or a cognitive impairment.

The open-ended questions in the interviews will explore how the organisation and adaption of services is accomplished in 1) the initial planning of services and home-visits carried out on the provider side after an order has arrived, and 2) in the daily planning of continued service organisation by service providers and how granted services (specialized and non-specialized to meet the needs of people with dementia) translate into delivered services/home visits (concerning staff continuity etc).

We will conduct digital interviews with 50 individual care providers, each representing a separate home care provision unit. A diversity in participating units will ensure representation of municipalities with varying size and geographical location. Appropriate interviewees (managers or care workers) with relevant knowledge will be invited to participate in the study based on a consultation with the managers of the units.

To provide specific data on the organisation of services including how certain decisions/orders on care services to people with dementia translates into certain combinations of services and visits of a certain number of care workers etcetera, the interviewees will also be asked to provide specific (anonymized) information regarding three care recipients with dementia with extensive care needs and services. The information regarding each person will include 1) The initial service order from the purchaser side of the municipality; 2) The individual implementation plan (*genomförandeplan*); and 3) How services were carried out a certain week (regarding staff continuity, frequency etcetera; see above). In addition, 20 of these 150 care recipients will also constitute the sample in sub-study 3.

The data will be analysed using a combination of analytical approaches to provide both in-depth knowledge and generalisable statistics on the organisation of social care services to people with dementia. The quantitative part of the data will be analysed using STATA 17 (StataCorp, College Station, TX) using common statistical methods, such as frequencies, proportions, Chi2, t-test, and Anova, depending on outcome variable and research question. The responses of the open-ended questions will be transcribed verbatim and analysed following the procedures of qualitative content analysis (26) (27).

##### Sub-study 3 – People living with dementia and their cohabiting partners receiving care services

The overall aim of sub-study 3 is to explore how services are performed in the direct meeting with older persons with dementia and how people living with dementia and their cohabiting partners experience and influence the performed care.

This study is conducted using 20 observations of regular care workers visits to the homes of care recipients, as well as 40 semi-structured interviews with the older persons with dementia and their cohabiting partners to document methods and practices in relation to experiences of the use and performance of the home care services. The sample consists of 20 care recipients selected from sub-study 2 as well as their cohabiting partners. The care recipients will be selected to ensure variation regarding municipalities that have specialized services for persons with dementia and municipalities that do not have specialized services. Concretely, this means that the researcher is present during the home visits on each observation occasion in the selected municipalities. The research group has vast experience of working with qualitative methods in different contexts and has specific competence for studying social relations -how people interact and relate to their surroundings. Interest is directed both towards general practices (how care and support are carried out) as part of people with dementia receiving care services, but also specific situations that may arise between care worker and the person with dementia.

The data collection method for the observations takes place primarily through field notes. Interviews with people with dementia and their cohabiting partners will be done during specific visits. The interviews are conducted with the aim of enriching the observations and increasing the understanding of the experience of the influence of the provision of and the content of the home care services. The interviews offer opportunities for the researchers to give feedback and ask about things and thoughts raised through the observations. The interviews thus fulfil the purpose of both providing new information and validating interpretations of previous information (28). All interviews are recorded with a Dictaphone, and then transcribed and anonymized. No personal data will be documented in writing.

The field notes from observations and interviews will be transcribed and analysed through qualitative content analysis (26) (27). Content analysis is useful when researchers want to avoid preconceived categories and instead allow the categories to emerge from the data (29). The combination of observations and interviews will make it possible to identify problems that arise in care practice and how the problems are handled by care workers in interaction with persons with dementia, as well as how the work is organized and communicated to the person with dementia and their cohabiting partners. Further, it will also be possible to capture the older persons living with dementia and their co-habiting partners own voices on how current services are experienced and how they want service interventions to be designed.

### Data management plans

#### Sub-study 1

The register database is stored on a secure server at Linköping University, accessible only to researchers involved in the project. All analyses are conducted directly on the server via remote access, and no data is transferred outside the server. The web surveys will also be downloaded and stored on this server.

#### Sub-studies 2 & 3

All data will be stored in secure cabinets at the Division of Ageing and Social Change (ASC), Linköping University. The material will be coded, and the code key/consent forms and data will be stored separately in different secure cabinets. The project leader will have access to the code keys and all data. All data will be pseudonymized, removing sensitive information such as personal data or any details that could reveal the identity of research participants. Data will be transferred to and stored in Linköping University’s high-security file vault. Each interview will be assigned a code, and only authorized researchers will have access to the code key, which will be stored on a separate server and in a secure cabinet. After the analysis is completed and scientific publications have been produced, the data will be stored for 10 years in the university’s file vault in accordance with university regulations. The code keys will continue to be stored on a separate secure server.

### Ethical considerations and declarations

The project was reviewed and approved by the Swedish Ethical Review Authority (Dnr 2024-05236-01). There are inherent risks associated with involving individuals with dementia and cognitive impairments as active participants in interviews and observations. In the early stages of dementia, many individuals retain the capacity to provide informed consent to participate in research. However, as the disease progresses, difficulties with comprehension, attention, memory, and communication may arise, making it challenging for researchers to determine whether consent can be considered truly informed.

We do not intend to select or exclude participants based on cognitive ability or impairment. However, we will take great care to ensure that all participants with dementia have understood the information about the project. Information will be provided both orally and in writing to all participants before consent is requested and given. For individuals with dementia receiving home care to participate, it is also required that their cohabiting family members provide consent—both for their own participation and for the participation of their relative with dementia.

Despite initial consent, there is a risk that participants may react negatively to the presence of researchers during interviews and/or observations. If the person with dementia—or their family member—shows signs of discomfort or distress in response to the researcher’s presence, the visit will be immediately terminated, and data collection in the home will cease.

The project is conducted by researchers with expertise in working with individuals with dementia, which is a strength in terms of preventing and managing such risks. While there are ethical challenges associated with involving people with dementia in research, it would also be ethically problematic to exclude them. The same applies to excluding individuals based on cognitive difficulties. To increase knowledge about the lived experiences of people with dementia and to ensure that individuals at all stages of the disease are included in conversations about their lives and the support they receive from elder care services, it is essential to include members of this group as participants in research projects such as this one.

The current lack of knowledge about how home care is organized, functions, is experienced, and adapted (or not adapted) for and by people living with dementia is problematic. It means that we also lack insight into the circumstances under which individuals with dementia, living in ordinary housing and receiving home care support, lead their lives. The project constitutes a review of today’s home care system, offering individuals with dementia and their cohabiting family members an opportunity to voice their experiences and perspectives on the support they receive. Our assessment is that the scientific and practical value of contributing such knowledge outweighs the low likelihood that participants experience discomfort from the observations, or from participating in interviews and sharing experiences with care organization.

### The status and timeline of the study

|  | **2025** |  | **2026** |  | **2027** |  |
| --- | --- | --- | --- | --- | --- | --- |
|  | *Spring* | *Fall* | *Spring* | *Fall* | *Spring* | *Fall* |
| **Substudy 1:** | - Planned survey data collection<br>- Developed web questionnaire | - First part of data collection of web questionnaire | - Finalizing establishing contact with needs assessors<br>- Finalizing the analyzes of web questionnaire<br>- Start analyzing register data | - Finalize analyzes<br>- Working on publications |  |  |
| **Substudy 2:** | - Planned datacollection<br>- Developed questionnaire<br>- | - Selected municipalities and care providers established contact to relevant interviewees<br>- conducted the internet-based interviews<br>- Recordings were sent for transcription |  | - Analyzing data<br>- Working on publications | - Finalize analyses<br>- Working on publications<br>- |  |
| **Substudy 3:** | - Planned observation scheme and developed | - Selected municipalities and care providers | - Finalize data collection | - Start analyzes<br>- Working on publications |  |  |
|  | semi-structured interview guide | - Started data collection | - Send material for transcription |  |  |  |
| **Project overall** | - | - | - | - |  | - Finalizing scientific paper<br>- Finalizing report<br>- Meeting with the reference group from Dementia Association |

As indicated in the table above, participant recruitment and data collection for the project as a whole are planned to be completed by the end of December 2026, with the first results anticipated around May 2027.

## Discussion

Mapping and describing social care services granted to people living with dementia involves two major challenges. The first challenge concerns the definition of the study population. People living with dementia constitute a heterogeneous group, ranging from newly diagnosed with mild symptoms to those in late stages of the disease, with severe care needs. In addition, not all individuals with dementia have a formal diagnosis. Social care services, according to SSA, are formally granted based on an assessment of individual needs, rather than a diagnosis. As, a result needs assessors – and home care providers – cannot be expected to know whether the older adults they assess or provide care to have a dementia diagnosis.

Hence, needs assessors and care providers interviewed about social care services for people with dementia may define the group differently or may not have information about a potential diagnosis. Such variation poses challenges for both data collection and analysis. To address this, the project applies the concept of “people living with dementia” in a pragmatic and analytical way, following approaches used in previous social scientific research (30). This means that the concept will not be applied as a strict diagnostic category but as a flexible construct acknowledging the diversity of the population and enabling comparisons across the three sub studies.

The second challenge is the lack of comprehensive national data on people living with dementia. Swedish registers most commonly used for information about dementia diagnosis-The Cause of Death Register and the National Patient Register (including inpatient and outpatient data)-have a very high specificity but only moderate sensitivity (31). This means that individuals recorded with a dementia diagnosis are very likely to truly have the condition, but a substantial proportion of people with dementia are not captured in these registers. The National register of Social Services also has limitations for research purposes. Despite its national coverage and improved data quality (32), there are still inconsistencies in reporting structures across municipalities. For instance, some municipalities report the number of granted home care hours, while others report only the type of service (6). Using only register data will not provide a complete picture of social care services granted to people with dementia, since no register will have full coverage of people living with dementia and all different types of home care services granted. In addition, qualitative data sources such as interviews with needs assessors and care providers are subject to methodological challenges. Local variations in how people with dementia are defined and understood may introduce selection and information bias, limiting the generalizability of the findings. Taken together, these limitations imply that neither register data nor interviews alone can provide a complete or fully valid picture of social care services for people with dementia. The strength of the project therefore lies in its methodological triangulation, combining register-based studies, surveys, observations, and interviews to enhance validity and robustness.

**Figure 1.**
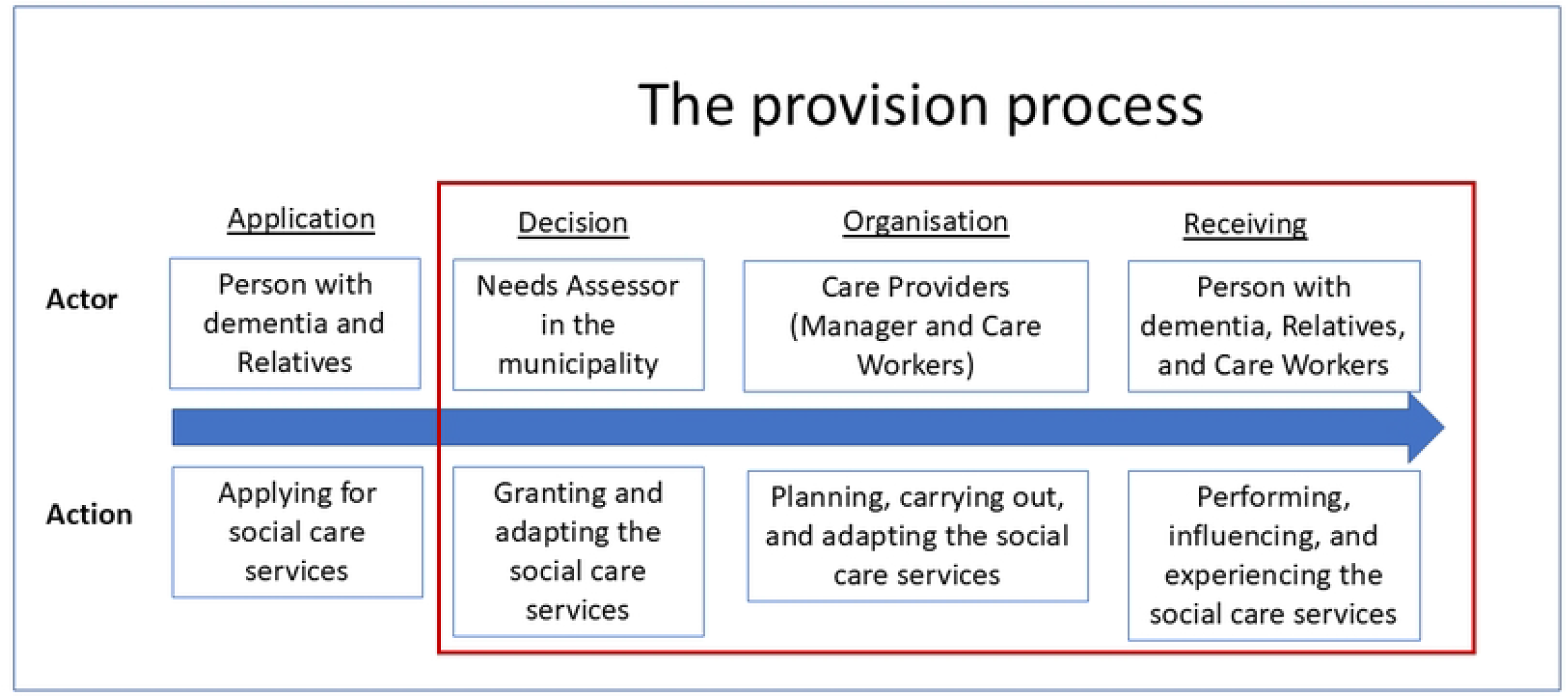

## Data Availability

There are no data associated with this protocol article. In the future, access to de-identified data for research purposes may be requested by contacting the authors.

## Authors’ contributions

Conceptualization: Annika Taghizadeh Larsson, Anna Olaison, Lars-Christer Hydén, Susanne Kelfve.

Funding acquisition: Annika Taghizadeh Larsson, Anna Olaison, Lars-Christer Hydén, Susanne Kelfve.

Project administration: Mattias Antar, Ceciel Heijkants, Jimmy Lindberg, Sofia Nordmark, Veronika Wallroth.

Writing – original draft: Annika Taghizadeh Larsson, Anna Olaison, Lars-Christer Hydén, Susanne Kelfve.

Writing – review & editing: Annika Taghizadeh Larsson, Anna Olaison, Lars-Christer Hydén, Susanne Kelfve, Mattias Antar, Ceciel Heijkants, Jimmy Lindberg, Sofia Nordmark, Veronika Wallroth.

## Supporting Information

None

## References

1. Odzakovic E, Hydén L-C, Festin K, Kullberg A. People diagnosed with dementia in Sweden: What type of home care services and housing are they granted? A cross-sectional study. Scand J Public Health. 2019;47(2):229–239.

2. Moberg L. Marketisation of Nordic eldercare: Is the model still universal? J Soc Policy. 2017;46(3):603–621.

3. Andersson K, Kvist E. The neoliberal turn and the marketization of care: The transformation of eldercare in Sweden. Eur J Womens Stud. 2015;22(3):274–287.

4. Blomberg S. Specialiserad biståndshandläggning inom den kommunala äldreomsorgen. Lund: Lund University; 2004.

5. Szebehely M. Vardagens organisering. Om vårdbiträden och gamla i hemtjänsten. Arkiv förlag; 1995.

6. Meinow B, Wastesson JW, Kåreholt I, Kelfve S. Long-term care use during the last 2 years of life in Sweden. J Am Med Dir Assoc. 2020;21(6):799–805.

7. Dunér, A. Equal or individual treatment: contesting storylines in needs assessment conversations within Swedish eldercare. Eur J Soc Work. 2018;21(4):546–558.

8. Olaison, A., Torres, S., & Forssell, E. Professional discretion and length of work experience: what findings from focus groups with care managers in elder care suggest. J Soc Work Pract. 2018;32(2):153–167.

9. Olaison, A., Knechtel, M., Torres, S., & Forssell, E. Dokumentationens roll för klientskapande processer i äldreinriktat socialt arbete: Spelar utlandsfödd bakgrund, kön och ålder någon roll? Sociologisk forskning. 2021;58(3):287–310.

10. Strandell, R. Care workers under pressure–A comparison of the work situation in Swedish home care 2005 and 2015. Health Soc Care Community. 2020;28(1):137–147.

11. Hammar LM, Alam M, Olsen M, Swall A, Boström AM. Being treated with respect and dignity? Perceptions of home care service among persons with dementia. J Am Med Dir Assoc. 2021;22(3):656–662.

12. McGovern J. Capturing the significance of place in the lived experience of dementia. Qual Soc Work. 2017;16(5):664–679.

13. Olsen M, Udo C, Boström A, Marmstål Hammar L. Important aspects of home care service: An interview study of persons with dementia. Dementia. 2021;20(5):1649–1663.

14. Polacsek M, Goh A, Malta S, Hallam B, Gahan L, Cooper C, et al. ‘I know they are not trained in dementia’: Addressing the need for specialist dementia training for home care workers. Heal Soc Care Community. 2020;28(2):475–84.

15. Wånell SE. Tillit om relationer: Om kvalitet i hemtjänsten - en kunskapsöversikt. 2015.

16. Brodaty H, Donkin M. Family caregivers of people with dementia. Dialogues Clin Neurosci. 2009;11(2):217–228.

17. Lethin C et al. Dementia care and service systems. BMC Health Serv Res. 2018;18:1–20.

18. McCabe M, You E, Tatangelo G. Hearing their voice: A systematic review of dementia family caregivers’ needs. Gerontologist. 2016;56(5):e70–e88.

19. Nilsson E. Framing dementia experiences in a positive light: Conversational practices in one couple living with dementia. Dementia. 2022;21(3):830–850.

20. Aaltonen MS, Martin-Matthews A, Pulkki JM, Eskola P, Jolanki OH. Experiences of people with memory disorders and their spouse carers on influencing formal care: “They ask my wife questions that they should ask me.” Dementia. 2021;20(7):2307–2322.

21. Sandberg L, Borell L, Rosenberg L. Risks as dilemmas for home care staff caring for persons with dementia. Aging Ment Health. 2021;25(9):1701–1708.

22. Morgan S. Positive risk-taking: A basis for good risk decision-making. Health Care Risk Rep. 2010;16(4).

23. Bowen ME, McKenzie B, Steis M, Rowe M. Prevalence of and antecedents to dementia-related missing incidents in the community. Dement Geriatr Cogn Disord. 2011;31(6):406–412.

24. Stevenson M, McDowell ME, Taylor BJ. Concepts for communication about risk in dementia care: A review of the literature. Dementia. 2018;17(3):359–390.

25. Sandberg L, Nilsson I, Rosenberg L, Borell L, Boström AM. Home care services for older clients with and without cognitive impairment in Sweden. Health Soc Care Community. 2019;27(1):139–150.

26. Spradley JP. Participant Observation. Waveland Press; 2016.

27. Lindgren BM, Lundman B, Graneheim UH. Abstraction and interpretation during the qualitative content analysis process. Int J Nurs Stud. 2020;108:103632.

28. Fejes A, Thornberg R. Handbok i kvalitativ analys. Liber; 2009.

29. Graneheim UH, Lundman B. Qualitative content analysis in nursing research: concepts, procedures and measures to achieve trustworthiness. Nurse Educ Today. 2004;24(2):105–112.

30. Palm R et al. People with dementia in nursing home research: a methodological review of the definition and identification of the study population. BMC Geriatr. 2016;16:78.

31. Rizzuto D, Feldman AL, Karlsson IK, Dahl Aslan AK, Gatz M, Pedersen NL. Detection of dementia cases in two Swedish health registers: A validation study. J Alzheimers Dis. 2018;61(4):1301–1310.

32. Meyer AC, Sandström G, Modig K. Nationwide data on home care and care home residence: presentation of the Swedish Social Service Register, its content and coverage. Scand J Public Health. 2022;50(7):946–958.

